# Estimating COVID-19 Cases on University Campuses Prior To Semester

**DOI:** 10.1101/2021.01.22.21250120

**Authors:** Alex Fout, Jude Bayham, Margaret J Gutilla, Bailey K Fosdick, Heather Pidcoke, Michael Kirby, Peter Jan van Leeuwen, Chuck Anderson

## Abstract

For many institutions of higher learning, the beginning of each semester is marked by a significant migration of young adults into the area. In the midst of the COVID19 pandemic, this presents an opportunity for active cases to be introduced into a community. Prior to the Fall 2020 semester, Colorado State University researchers combined student home locations with recent case counts compiled by the New York Times to assign a probability to each individual of arriving with COVID19. These probabilities were combined to estimate that there would be 7.8 new cases among the on-campus population. Comprehensive testing of arriving students revealed 7 new cases, which validated the approach. The procedure was repeated to explore what could happen if students had returned to campus after Fall break. The estimate of 48 cases corroborated the University’s early decision to transition to fully remote learning after break.

## Introduction

Colleges and universities were hot spots for COVID-19 during the Fall Semester 2020 (*1*). While public health officials can continuously monitor recent case rates to monitor the local epidemic, the commencement of the university semester causes a migration of young adults into the area, often from geographically diverse home locations. Since COVID-19 prevalence varies considerably across the country, this migration poses a potential risk to students, faculty & staff, and the broader community (*2*). An accurate estimate of the number of COVID-19 cases arriving to campus can greatly inform the development of public health strategies, such as initial quarantine space needs, contact tracing demands, and ensure proper initialization in epidemiological models. The objective of this study was to use student home county and county disease prevalence estimates to determine the expected number of COVID-19 cases upon students arriving on campus. This method is demonstrated using data from Colorado State University’s (CSU) on-campus student resident population and comprehensive testing results that occurred in the week prior to the start of Fall 2020 classes. The estimated prevalence (0.16%) agreed with the observed prevalence (0.17%) obtained from CSU pre-semester testing. This analysis was repeated to assess the risk of students returning to campus post-Thanksgiving break, and the large number of estimated cases corroborates the University’s decision to move classes fully online after break.

## Methods

Colorado State University is a Public Land Grant University in Fort Collins, Colorado with approximately 27,000 undergraduate, 8,000 postgraduate students. International students comprise roughly 2,000 of the overall student population.

The home state and county of students was used to estimate a likelihood of COVID-19 infection. The estimation procedure assigns a probability of testing positive to each incoming student based on their home location, then aggregates the probabilities to produce an estimated number of cases. University records indicated home residence ZIP codes for US students arriving to campus in August 2020. The probabilities were estimated by dividing the number of recent cases by the total population in a student’s home location. The number of “recent cases’’ was taken to be the change in total New York Times reported COVID-19 cases (*3*) in a 14 day window, to coincide with the upper estimate of the viral incubation period. Population counts were derived from the 2019 American Community Survey (5 Year) (*4*). ZIP Codes were mapped to counties using ZIP files from the US Department of Housing and Urban Development (*5*). Cases and population were aggregated at the county and state level to produce estimates at different resolutions (i.e. assigning a probability based on a student’s home state and county, or just their home state). Given these probabilities, the number of arriving cases is a sum of Bernoulli experiments (assumed independent), so the expected number of cases is the sum of the probabilities. To estimate the variability in cases, the infection status for each student was simulated using their respective probability, the results were summed to get a count of infected individuals. Repeating this process 10,000 times produced a distribution of case counts, from which 95% confidence intervals were obtained.

For comparison, this estimation procedure was repeated by assuming a uniform probability of infection for all students, based on the case counts and populations in either the entire US, Colorado, or Larimer County (the home county of CSU). The US based estimates show the impact of “individualizing’’ a student’s probability based on their home location, and the Colorado & Larimer County estimates shed light on the differential risk introduced by the incoming students, compared to an equivalent population from only the state or county.

For external validation, estimates were compared against CSU’s comprehensive testing of incoming students prior to the start of the Fall 2020 semester. The confirmatory test was an FDA Emergency Use Authorized digital droplet PCR assay from Biodesix Inc.

Finally, the estimation was repeated for the hypothetical scenario where students returned to campus after fall break. These estimates did not account for reported cases during the semester and thus assume everyone would have been both returning and susceptible. Pre-semester estimates used case counts from 24 July - 6 August 2020, and post Fall Break estimates used case counts from 16 - 29 November 2020. This project was reviewed and approved by CSU’s Institutional Review Board under protocol 20-10414H.

## Results

Table 1 summarizes the case estimates using individualized probabilities of infection based on student home county and home state, and the comparison estimates using constant probabilities based on case counts from the entire US, Colorado only, or Larimer County only, as well as ground truth for the pre-Fall semester estimates. Figure 1 depicts maps of where students originate and the relative case rates for different regions (US states and Colorado counties).

**Table 1:**
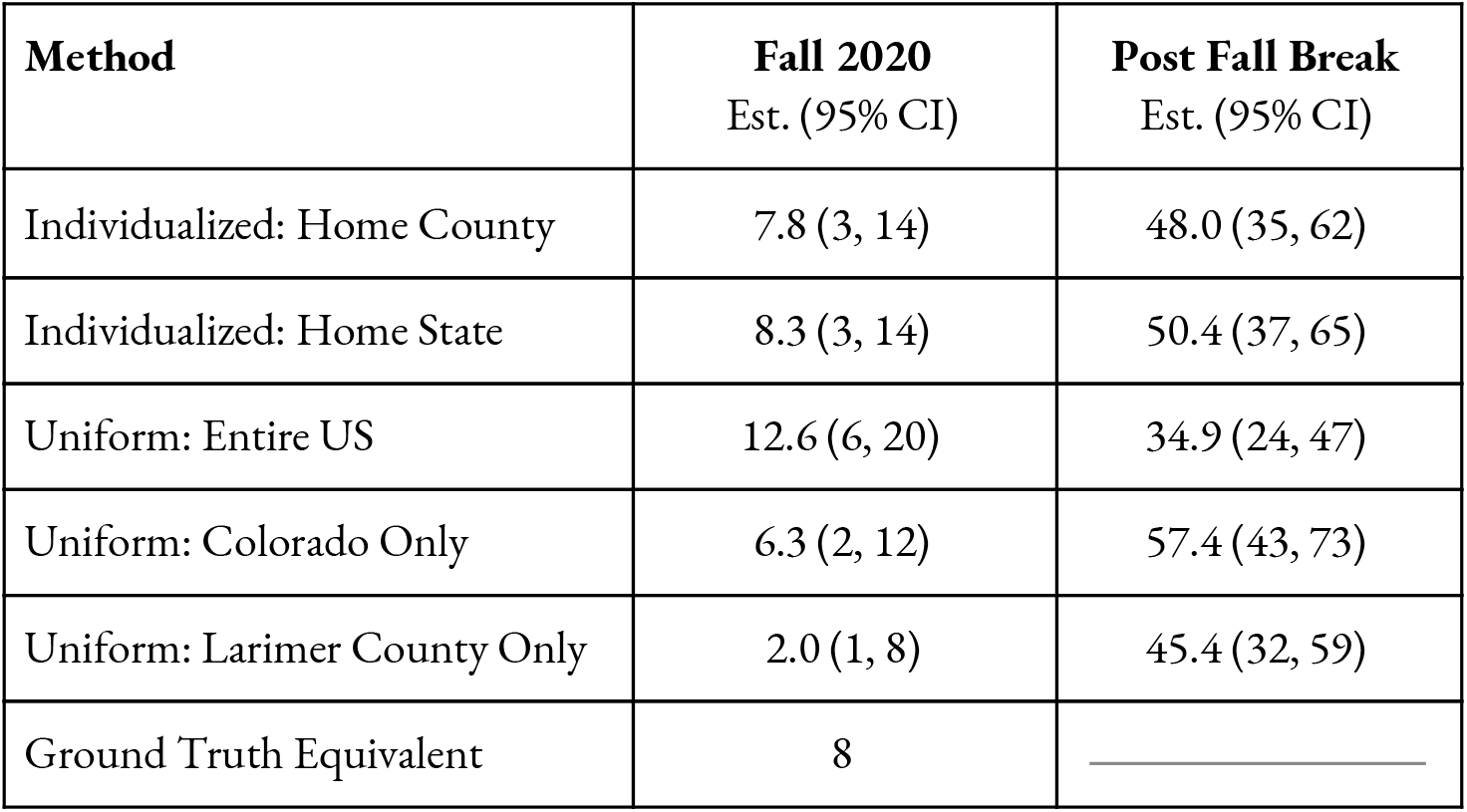
Estimated student COVID-19 cases before the start of Fall semester, and after fall break based on different estimates of the probability of infection for each of 4,906 on-campus resident students. Because the university conducted comprehensive testing of incoming students prior to the Fall semester, ground truth is known for that period. Out of 4,046 tests, CSU reported 7 positive results. This equates to roughly 8 cases in 4,906 students (for comparison).

**Figure 1:**
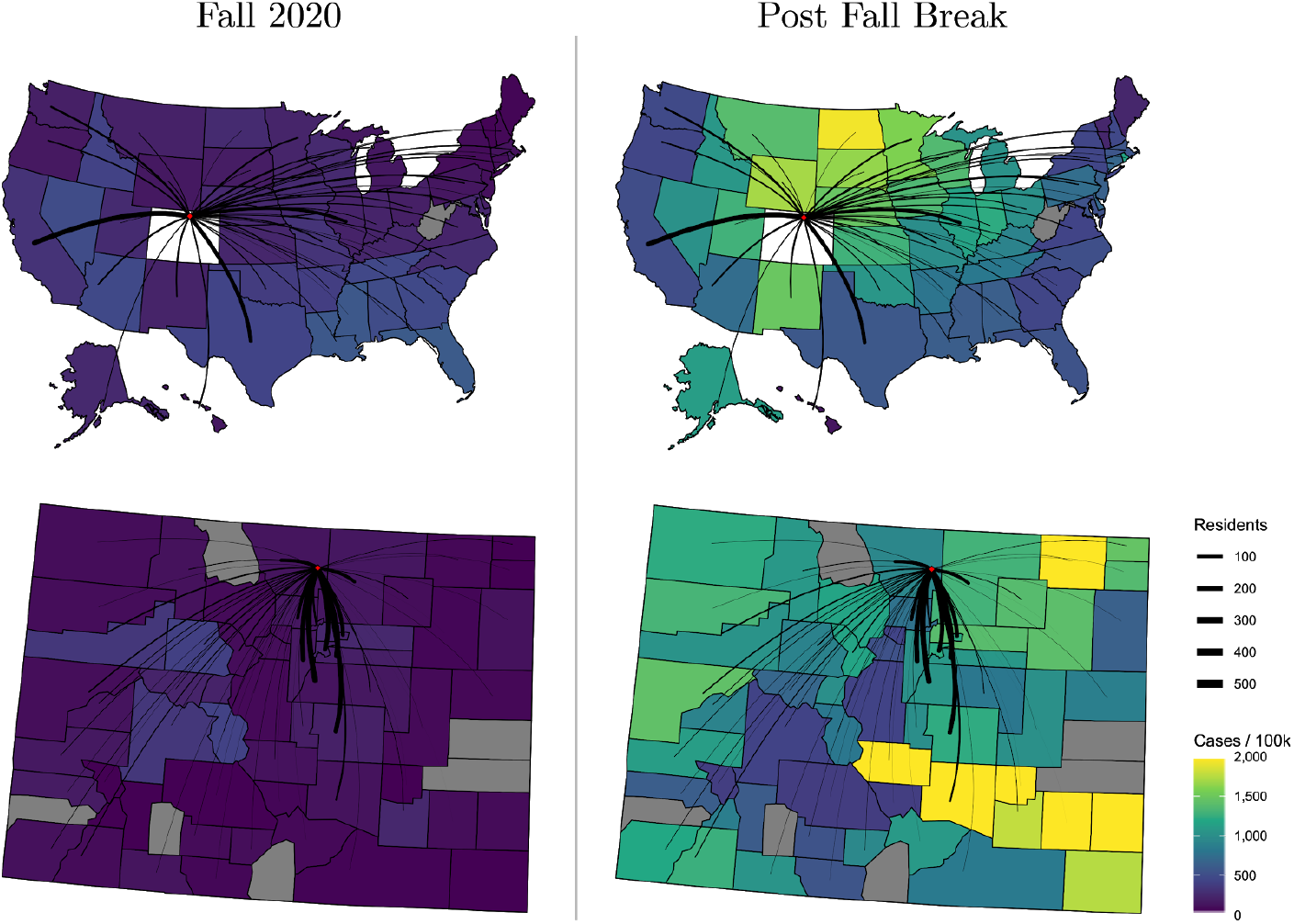
Maps of US States and Colorado counties illustrating the residence location for CSU Fall 2020 campus residents. Lines are sized in proportion to the number of students originating from that location, and the locations are shaded by the recent COVID-19 case rates. The left maps reflect case rates from 24 July - 6 August 2020 just prior to the Fall 2020 semester; the right maps reflect the period 16 - 29 November 2020 over Fall break. The red dot indicates CSU.

The individualized estimates using home county and home state generally agree for both time periods, in both the point estimates and the simulated confidence intervals, suggesting that heterogeneity across states is more important than heterogeneity within a state. These estimates differ more strongly from the uniform estimates showing the impact of incorporating home location.

## Discussion

Geographic diversity has been a hallmark of COVID-19 prevalence in the US. Most students traveling to Colorado State University come from counties in Colorado, so the estimates are mostly governed by the recent cases in the state. However, the effect of students from out of state is noticeable in both the pre-Fall semester and post Fall break periods. Indeed, the individualized locale-specific point estimates and confidence intervals appear to split the difference between the entire US and Colorado only estimates. The key public health implication is that in the pre-Fall semester time period, higher case rates outside the state suggest an increased risk from out of state students. Yes, the situation was reversed in the post-Fall break environment, where Colorado was experiencing an active outbreak, and out-of-state students posed comparatively lower risk.

During comprehensive testing prior to Fall semester, 7 out of 4,046 students tested positive, which adjusts to 8 in 4,906, which is very close to the locale-specific estimates. In terms of prevalence, the “Individualized: Home County” method gives 0.16%, compared to the ground truth prevalence of 0.17%.

By virtue of being simple, this estimation method should be accessible to other institutions of higher education and other organizations which draw a geographically diverse audience. Some possible examples of entities beyond educational institutions that might benefit from this exercise include airports, ski areas, resorts, and other travel destinations. The practicability of this method is tied to the ability to identify the “home location’’ of an individual, which should already be known by an airport, for example. Importantly, no other personally identifying information is needed. Reliable case counts and populations are publicly available for many locations. This method could also incorporate model based projections of case counts and visiting individuals in order to estimate future cases for planning purposes.

Many factors influence the likelihood of an incoming student testing positive which we are not accounted for by this method, such as behavioral patterns, living arrangement, and family structure. However this information is rarely available, and is more sensitive in nature than place of origin.

Ascribing a probability to each student based on recent cases, implicitly assumes homogeneity in the respective local populations, however the differences within each home location are assumed less important than the differences between different home locations. Incoming students are generally of similar age (roughly 18-21), so any age-specific adjustments to infection rates are assumed consistent across all home locations.

Infections are assumed independent of one another, which is reasonable for the vast majority of incoming students (with the exception of, for example, two students from the same family). This analysis excludes international students due to the varying reliability of case counts globally, concern over comparing case counts reported by different countries, and the low number of students arriving from outside the US. It is also possible that reported case counts will underestimate the true number of infections, which is not accounted for here.

## Data Availability

Student zip codes are not releasable, and other data sources are publicly accessible.

https://github.com/nytimes/covid-19-data

https://www.huduser.gov/portal/datasets/usps_crosswalk.html

https://www2.census.gov/programs-surveys/acs/summary_file/2019/data/5_year_entire_sf/Tracts_Block_Groups_Only.zip

## Acknowledgements

Laura Jensen, Colorado State University; Cory Hudson, Colorado State University, Alan Rudolph, Colorado State University, Erin Staples, Centers for Disease Control

Funding was provided by the Office of Vice President for Research, Colorado State University

## Summary

### What is already known about this topic?

Geographic heterogeneity in COVID-19 prevalence paired with human mobility drive changing infection rates. College towns and campuses are susceptible to these drivers as large numbers of young adults travel to school at the beginning of an academic term.

### What is added by this report?

Colorado State University estimated the number of expected cases in their on-campus resident population at the beginning of the Fall 2020 semester using information about the case rates of student home locations. These estimates differ from those assuming all students have a uniform probability of infection that is reflective of the national, state or local county case rates.

### What are the implications for public health practice?

Large shifts in a local population can cause dramatic changes in infection rates and possibly overwhelm local health support services, such as testing, quarantine capacity, and contact tracing. Accurately estimating the number of expected cases associated with an influx into an area can allow Universities to adequately prepare.

